# Investigation of air change rate and aerosol behavior during an outbreak of COVID-19 in a geriatric care facility

**DOI:** 10.1101/2022.01.27.22269512

**Authors:** Yo Ishigaki, Shinji Yokogawa, Yuki Minamoto, Akira Saito, Hiroko Kitamura, Yuto Kawauchi

## Abstract

**Background:** Ventilation plays an important role in controlling aerosol transmission of coronavirus disease (COVID-19), and mass transmission of COVID- 19 has been reported in poorly ventilated areas.

**Objective:** A real-world mass infection outbreak which occurred in an elderly nursing home, in Miyagi Prefecture, Japan, was simulated experimentally and numerically to investigate the controlling factors and quantify the effectiveness of various natural ventilation settings by means of air change rate (ACR).

**Methods:** Using the CO_2_ tracer gas method, the ACR values at the time of the outbreak were estimated to be 2.0–6.2 h^-1^ in rooms in the facility. Furthermore, a low-cost intervention of opening windows improved the ventilation frequency by a factor of 1.48–5.74. This implies that advective fluid flows are the key in the spread of high CO_2_ concentration zones. A numerical simulation was performed to obtain spatio-temporal evolution on such high CO_2_ concentration zones under similar conditions to the present experiment.

**Results:** The results showed that ventilation was significantly dependent on the window opening conditions in all rooms (p-values ranging from 0.001 to 0.03 for all the rooms). In contrast, there was no significant dependence on the location of the sensor in any of the areas. Development of high CO_2_ concentration zones occurs in the first few minutes. Furthermore, the leading edge of such zones towards the common room yields a relatively high fluid velocity, suggesting that the large-scale advective flow dictates the spread of such high CO_2_ concentration zones.

**Conclusions:** The present results suggest that secondary infections could occur due to the aerosol advection driven by such large-scale flows, even when the building design adheres the ventilation guidelines. In elderly care facilities, open architectural spaces are recommended to realize quality of life and monitor residents. However, management is required to reduce the downwind infection risk from aerosols and ACR.

## Introduction

Ventilation plays an important role in controlling aerosol transmission of coronavirus disease (COVID-19), and mass transmission of COVID-19 has been reported in poorly ventilated areas [1, 2]. Menzies et al. [3] analyzed determinants of the risk of secondary transmission of tuberculosis and measles in various hospital settings and reported that an average of less than two ventilation cycles per hour (air change rate per hour; ACR) in the examination room was significantly associated with tuberculin conversion. Based on this study, the Centers for Disease Control and Prevention (CDC) has set a standard for ventilation in negative pressure rooms for the isolation of patients with infectious diseases, with an ACR recommendation of six (for existing buildings) to twelve (for new buildings) with a safety factor [4]. The World Health Organization (WHO) has set a standard of 576 m^3^/h per person for natural ventilation in health facilities dealing with infectious diseases, based on the CDC standard of twelve ACR, assuming that one patient occupies a space of 4 × 2 × 3 m^3^, and with a doubled safety factor [5]. In Japan, the Ministry of Health, Labor and Welfare (MHLW) has suggested a similar value of 576 m^3^/h per person for health facilities. Further, the MHLW, referring to the above-mentioned guidelines from other countries, has recommended a ventilation rate of 30 m^3^/h^-1^ per person in general commercial facilities to deter indoor aerosol transmission of COVID-19 [6].

Peng et al. [7] attempted to explain the risk of COVID-19 aerosol infection considering ACR and indoor carbon dioxide concentrations; however, comprehensive epidemiological studies with large-scale validation are needed to quantify these relationships. Therefore, it is important to conduct field surveys on the ventilation performance and aerosol behaviors at sites where mass infections of COVID-19 have occurred.

In Japan, there has been a series of outbreaks of COVID-19 in nursing facilities. However, no study has been carried out with on-site investigation using ACR and indoor CO_2_. In the present study, an outbreak site of severe acute respiratory syndrome coronavirus 2 (SARS-CoV-2) in a nursing home in Miyagi Prefecture, Japan was investigated to measure the ACR and estimate the ventilation volume using a CO_2_ tracer gas method at various window opening conditions. In addition, the aerosol distribution was analyzed via computational fluid dynamics to determine the three-dimensional aerosol behavior. The scope of the present study was to find causative factors of an actual outbreak of mass infection in an elderly care facility from the perspective of ACR and aerosols and discuss recommendations for preventing recurrence.

## Methods

The elderly care facility investigated in the present study is located in Miyagi Prefecture, Japan, where a total of 59 cases were reported in the same building by April 2021. Of these, 36 were users of the facility (including both residents and daily visitors) and the other 23 were facility staff members. After the outbreak, the Disaster Medical Assistance Team (DMAT) was dispatched for zoning, and critically ill patients were hospitalized, after which the infection situation was in control. The field survey reported here was conducted in August 2021.

Based on interviews with the facility staff, five separate areas in the building, where large-scale secondary infection occurred, were extensively studied: (a) regular bathroom, (b) nursing bathroom, (c) shared room, (d) private room, and (e) day room, as shown in Figure 1. Both regular and nursing bathrooms are used by the residents, and staff members accompany them for assistance. In general, the risk of aerosol infection is considered to increase in the bathrooms because neither the residents nor the staff wear a mask because of the high humidity environment. Furthermore, a care recipient using the nursing bathroom requires high-level care, and the staff must talk to them while making contact, which is expected to increase the risk of transmission. In the shared room, there were a maximum of four beds in the room, three of which were for residents, and the other bed was unused at the time of the outbreak. Although the residents were instructed to wear masks at all times, it was difficult to enforce on patients with certain conditions such as dementia. A private room is a room for a single resident; therefore, the risk of aerosol infection is relatively low. It is segregated from the corridor by a curtain. Notably, the first resident who tested COVID positive was found in a private room next to the day room; then, the mass infection outbreak occurred. The day room is a place for relaxation and has free access for all residents and other visitors temporarily visiting the facility between 6:00 a.m. and 8:00 p.m. daily, and it is furnished with chairs, tables, and televisions. Since the day room is frequently used by multiple people including care recipients and staff, the risk of infection is expected to be relatively high. These rooms were first investigated individually in the primary measurements described later. Therefore, the layout of these rooms in the entire facility floor plan is irrelevant. Furthermore, there were several transmission routes involved in the studied mass infection outbreak event. The present study focuses on airborne transmission via aerosols; however, other transmission routes, such as direct contact, were not excluded.

**Figure 1.**
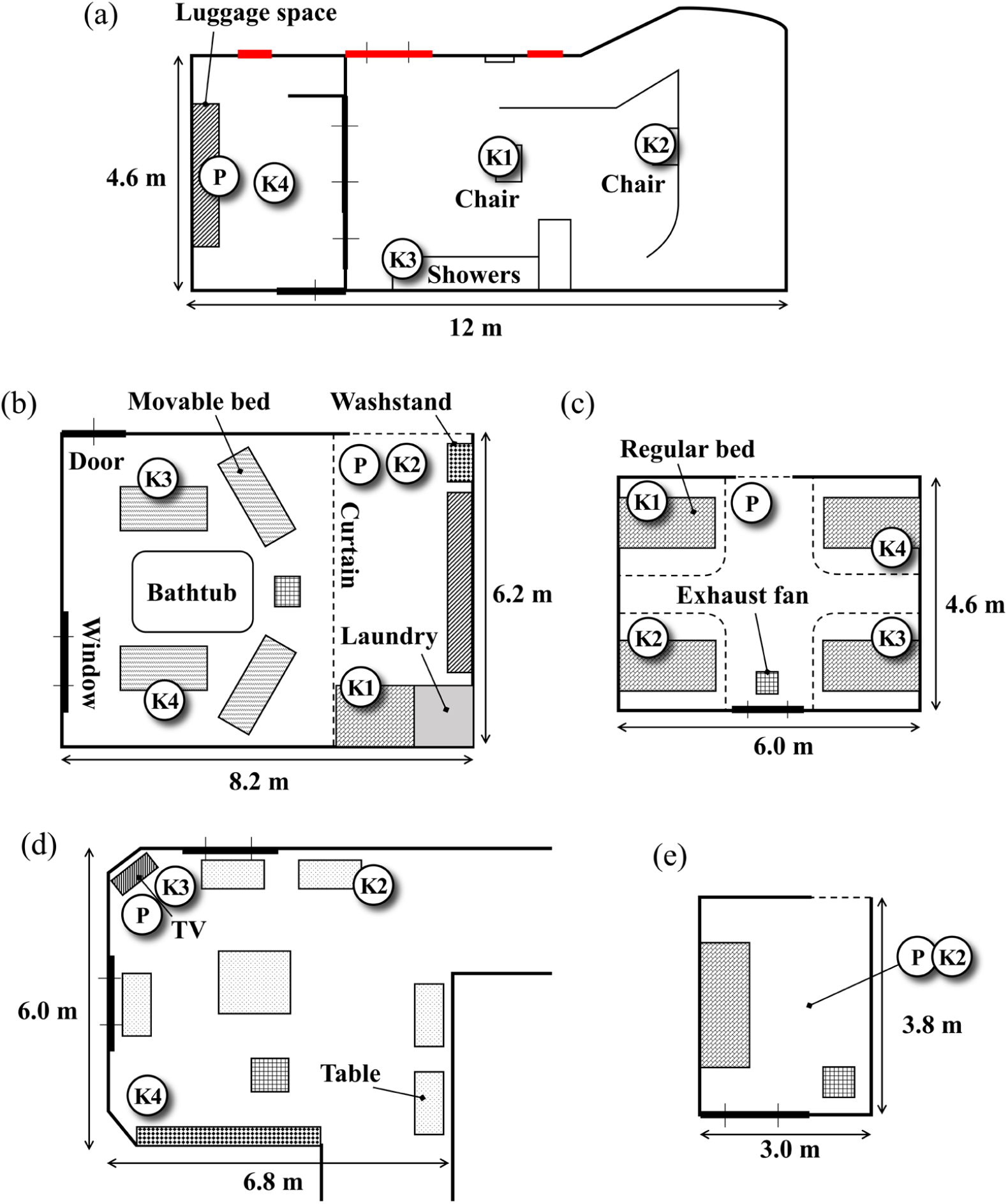
Floor plan of each room investigated in the primary measurement. Locations of different sensors, P, K1—K4, are also shown. (a) Regular bathroom, (b) nursing bathroom, (c) shared room, (d) day room, and (e) private room. Ceiling heights of these rooms were 2.6, 2.4, 2.7, 2.4, and 2.4, respectively.

## Measurements

The on-site measurement was conducted under the guidance of an industrial physician to ensure safety, while confirming that the polymerase chain reaction (PCR) test results of all residents, staff, and researchers were negative, and safety measures, such as wearing personal protective equipment and disinfection, were taken. In the present study, two types of experiments were performed, namely primary and secondary measurements.

### Primary Measurement

In the primary experiment, the tracer gas method was employed to measure the ACR in the five areas as discussed. CO_2_ was used as a tracer gas, which was obtained by vaporizing dry ice in the study room. Two types of Non-Dispersive-Infrared (NDIR)-type CO_2_ sensors ere used. The first was a mobile CO_2_ sensor (Yaguchi Electric Corporation, Miyagi, Japan) equipped with an NDIR sensor SCD-30 (Sensirion AG, Stäfa, Switzerland). Any measurement performed with this sensor is hereafter named “P”. The second sensor was a TR-76Ui (T&D Corporation, Nagano, Japan) and is named “K” with an index of measurement locations. A total of five sensors (one P and four K sensors, K1–K4) were used during the measurements. Figure 1 shows the arrangement of these sensors in each room. All sensors were placed at a height of approximately 1 m from the ground.

The measurements were conducted in each room as follows:

1. The CO_2_ sensors were installed at the locations shown in Figure 1 and measurement started.
2. The mechanical ventilation system in the room of interest was turned off, and the windows and doors were closed, if there were any, to create a closed room.
3. Dry ice was placed in the room of interest to achieve a CO_2_ concentration which was sufficiently high compared to the background (approximately 400 ppm). The concentration should be at least 2000 ppm but must not exceed the permissible concentration of 5000 ppm (at eight hours of exposure) specified by the Japanese Industrial Safety and Health Law. In addition, a blower was used to sufficiently stir the room air with the generated CO_2_ gas because the gas evaporated from the dry ice yields a low-temperature and tends to remain close to the floor.
4. When the CO_2_ concentration sufficiently increased, the dry ice was removed from the room, and the mechanical ventilation system and conditions of windows and doors were set according to the target measurement conditions. This time is denoted as *t*_0_.
5. The room was immediately vacated to avoid CO_2_ addition from breathing, and this time was defined as the start of ventilation. This time is denoted as *t*_*sta*_.
6. The CO_2_ concentration was remotely monitored from outside the room until the concentration decreased sufficiently.
7. Once the CO_2_ concentration decreased sufficiently, the measurement was finished, and this time is denoted as *t*_*end*_. The time-series measurement data from *t*_*sta*_to *t*_*end*_ were saved for analysis and used to estimate the ACR.
8. If the CO_2_ concentration was still sufficiently high, step 3 could be omitted, and we could proceed to Step (4).

The measured CO_2_ concentration data were processed to calculate the ACR for each room. The Seidel equation is:

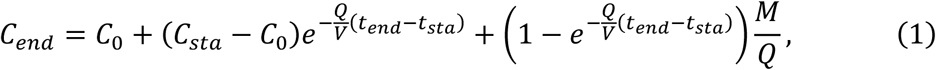

where *C*_*end*_ is the CO_2_ concentration (ppm) at *t* = *t*_*end*_, *C*_0_ is the steady state value of the CO_2_ concentration when there is no pollution (ppm), *C*_*sta*_ is the concentration (ppm) at *t* = *t*_*sta*_, *V* is the volume of the room (m^3^), *Q* is the ventilation rate (m^3^/h), and *M* is the rate of pollutant generation (ppm·m^3^/h). Clearly, the measurement start and end times, *t*_*sta*_ and *t*_*end*_, in the equation are in hours. Furthermore, in the present study, *C*_0_ was assumed to be 400 ppm. Since the room was vacant during the measurement, *M* = 0 in Equation (1). This leads to:

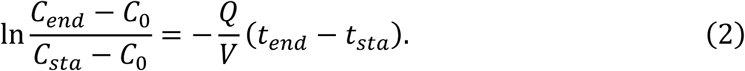

The ACR value, calculated as *Q*/*V* (1/h), was the decrease in CO_2_ concentration from *t*_*sta*_ until *t*_*end*_.

Notice that the next Heading Style (Heading Style 4 in this case) is used. Click on the different headings to see their Heading Style in the “Home” ribbon under “Styles”. Always have at least 2 of the same subheading level in a section.

### Secondary Measurement

In addition to measuring the five rooms individually, secondary measurement was performed, as shown in Figure 2. This secondary measurement was to investigate the effect of interplay between air flows in the private and day rooms because these rooms are spatially connected via a 2–3 m corridor. This fluid dynamical aspect suggests the possibility that aerosol leaked from the private room to the day room, and a resident who was COVID-positive in the early stage of the mass outbreak was isolated in the private room, as described in the earlier section. For the secondary measurement, the sensors were placed as shown in Figure 2, the private room was filled with CO_2_ gas, measurement steps (1)-(6) were followed, and gas leakage from the private room to the day room was investigated.

**Figure 2.**
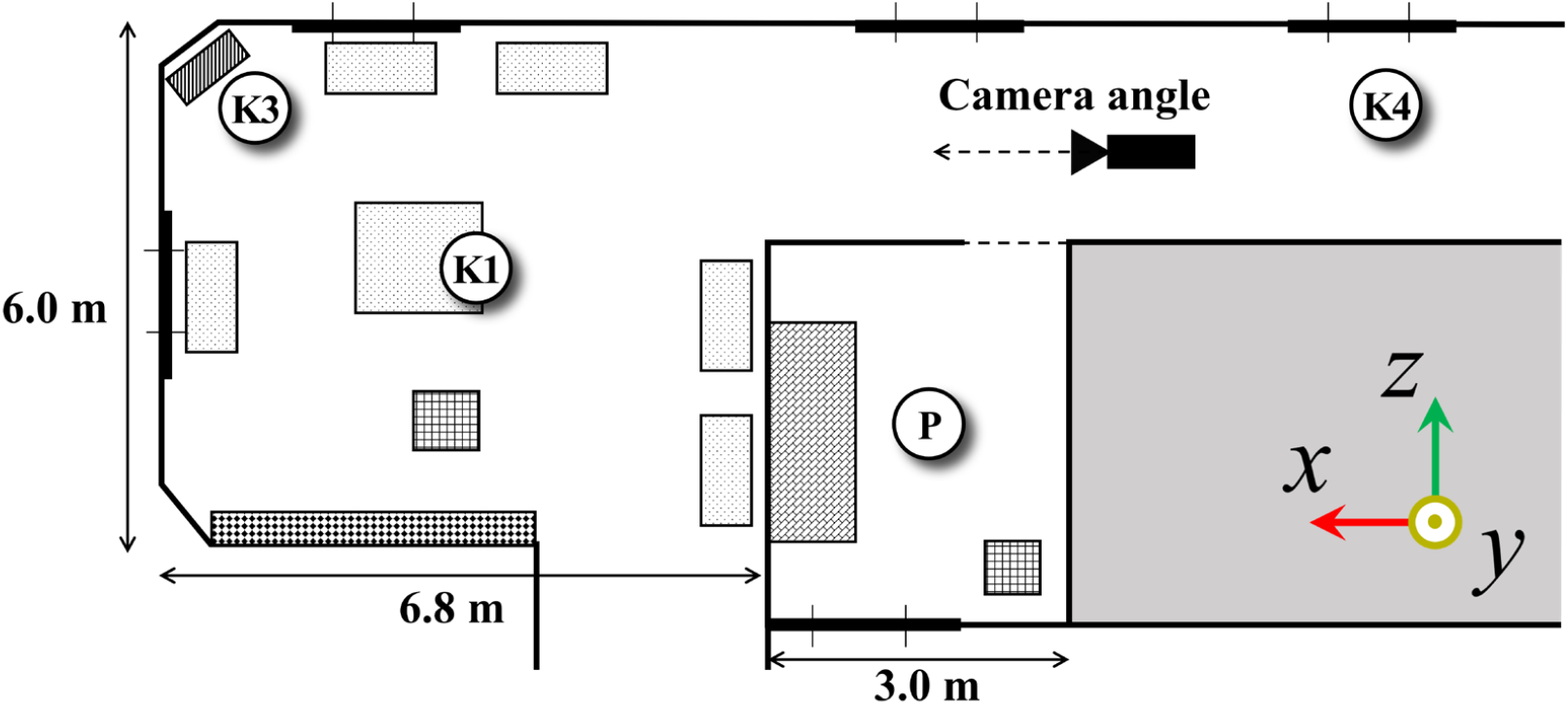
Floor plan including the private and day rooms connected by the short corridor, and sensor locations for the secondary measurement. Orientation axes are shown for the numerical simulation. The first COVID-positive resident was found in a private room next to the day room.

Figure 3 shows a direct photograph of the CO_2_ smoke which is leaking from the private room shown in Figure 2 to the corridor leading to the day room, taken from the camera angle shown in Figure 2. A substantial amount of CO_2_ smoke leaked from the gap between the curtain and the floor and advected towards the ceiling. By the time it reached the day room, a high concentration of CO_2_ smoke was observed near the head height of a person sitting in a wheelchair.

**Figure 3.**
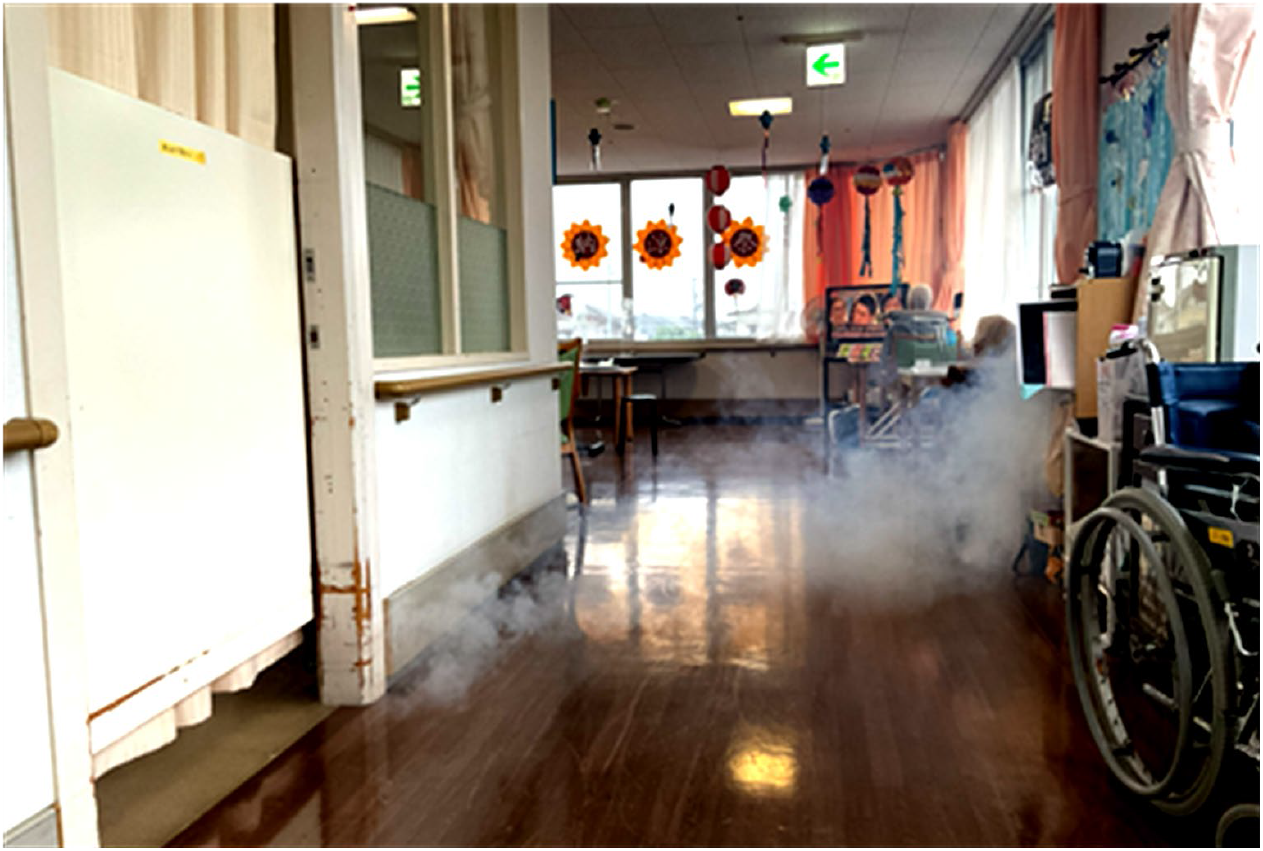
Direct photograph of the CO_2_ smoke leaking from the private room with the first COVID-positive patient into the corridor leading to the day room with multiple lounging tenants (taken from the camera angle shown in Figure 2).

## Numerical Simulation

Because spatio-temporal CO_2_ concentration distribution is not easily obtained from the experiment, a numerical simulation of gas leakage from the private to the day rooms was performed. This simulation corresponds to the secondary measurement. The simulation was performed using Flowsquare+ (version 2021R1.0, Nora Scientific Co. Ltd., Kanagawa, Japan) which solves transport equations for mass (density, *ρ*):

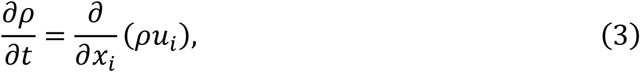

for momentum *ρu*_*i*_:

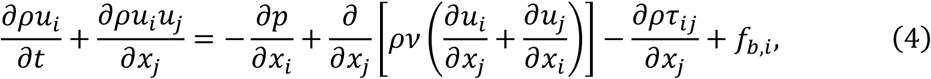

for energy (temperature, *T*):

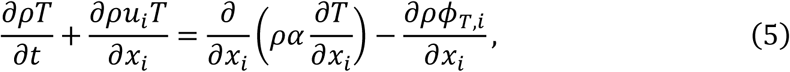

and for mass fraction of CO_2_ gas generated by the dry ice *Y*_*CC*2_:

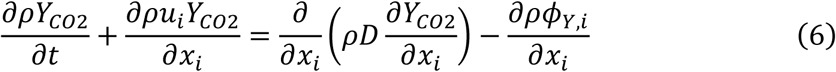

in a large eddy simulation (LES) context. Subscript indices *i* =1, 2, and 3 correspond to the directions *x, y*, and *z*, respectively. *ν, α*, and *D* are kinematic viscosity, thermal diffusivity, and molecular diffusivity, respectively, and *ρ ν* = 2.0 × 10^−6^ (kg/m/s), *α* = *ν* /*Pr* and *D* = *ν* /*Sc*, where the Prandtl number (*Pr*) = 0.7 and the unity Schmidt number are considered. The subgrid-scale (SGS) stress tensor, *τ*_*ij*_, and scalar fluxes, *ϕ*_*T,i*_ and *ϕ*_*Y,i*_, were modelled based on static Smagorinsky and gradient diffusion models, respectively. The transport equations were discretized onto *N*_*x*_ × *N*_*y*_ × *N*_*z*_ = 175 × 90 × 30 uniform mesh points using a second order finite difference scheme in space and advanced in time using the explicit Euler method. Only the advection term was calculated using a first order upwind scheme. The domain dimensions were *L*_*x*_ × *L*_*y*_ × *L*_*z*_ = 17.5 × 3.0 × 9.0 (m^3^). The physical boundaries which did not coincide with the Cartesian mesh were expressed by a second order immersed boundary method.

The fluid in the entire domain was initialized at 18 °C and *Y*_*CC*2_ = 0.0. The warm air fed from the room AC yielded a temperature of 30 °C. The fluid velocity issued from the air conditioner installed in the private room was *u*_*x*_, *u*_*y*_, *u*_*z*_ = (0, -1, 2) m/s, while that of the air conditioner installed in the day room was (0, -1, 1) m/s. The ventilation fan in the private room was turned off, while the ventilation fan in the day room discharged the room air at a velocity of (0, 10, 0) m/s. These settings were based on the measurement conditions considering the situation during mass infection outbreaks. To mimic the aerosol dispersion from the COVID-19 positive person in the private room, a small inflow boundary was considered on the bed with a constant (CO_2_) gas flow at 36 °C at a velocity of (0, 0.707, 0.707) m/s; therefore, its magnitude was 1.0 m/s. Furthermore, the fluid flow issued at this inflow boundary yield *Y*_*CC*2_ = 1.0.

During the numerical simulation, measurement probes were located for P, K1, and K3 as with the measurements, as shown in Figure 2, which facilitates the validation of the simulation results by comparing the measured data. The measurement at K4 was not performed for the simulation because of the close proximity to the computational boundary.

## Results

Table 1 summarizes the ACR values calculated based on Equation (2) from the measured CO_2_ concentration with each sensor in each room introduced in Figure 1. Here, “window opening condition” refers to the state of window opening and closing that was set in step (4), as discussed in Method section. For example, for the regular bathroom, both windows in the bathrooms and corridor were closed when a group outbreak occurred. In Table 1, values in the “Ave.” column are the arithmetic mean of the calculated ACR values recorded by different sensors in each room. The ratio, *r*_*ACR*_, denotes the ratio of the ACR values between “Open” and “Close” conditions. Thus, this ratio quantifies how much the ventilation volume improved by opening window(s). Furthermore, the values in the “Ventilation volume” column show the ventilation volume per person per hour calculated by multiplying the average ACR by the volume of the room and dividing it by the room capacity. As indicated in the Introduction, the MHLW requires at least 30 m^3^/h per person in general commercial facilities [6], the CDC requires at least six ventilation cycles per hour in negative pressure rooms (existing buildings) [4], and the WHO requires at least 576 m^3^/h per person in health facilities [5]. If the measured hourly ventilation volume complies with these MHLW, CDC, and WHO standards, the value was marked with †, ‡, and §, respectively.

**Table 1.**
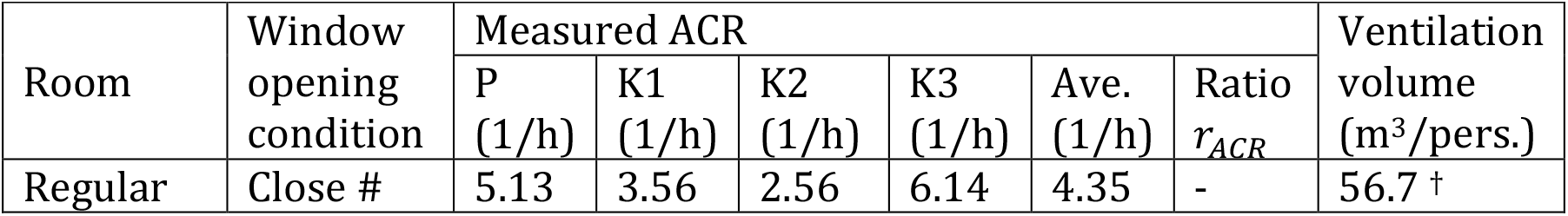
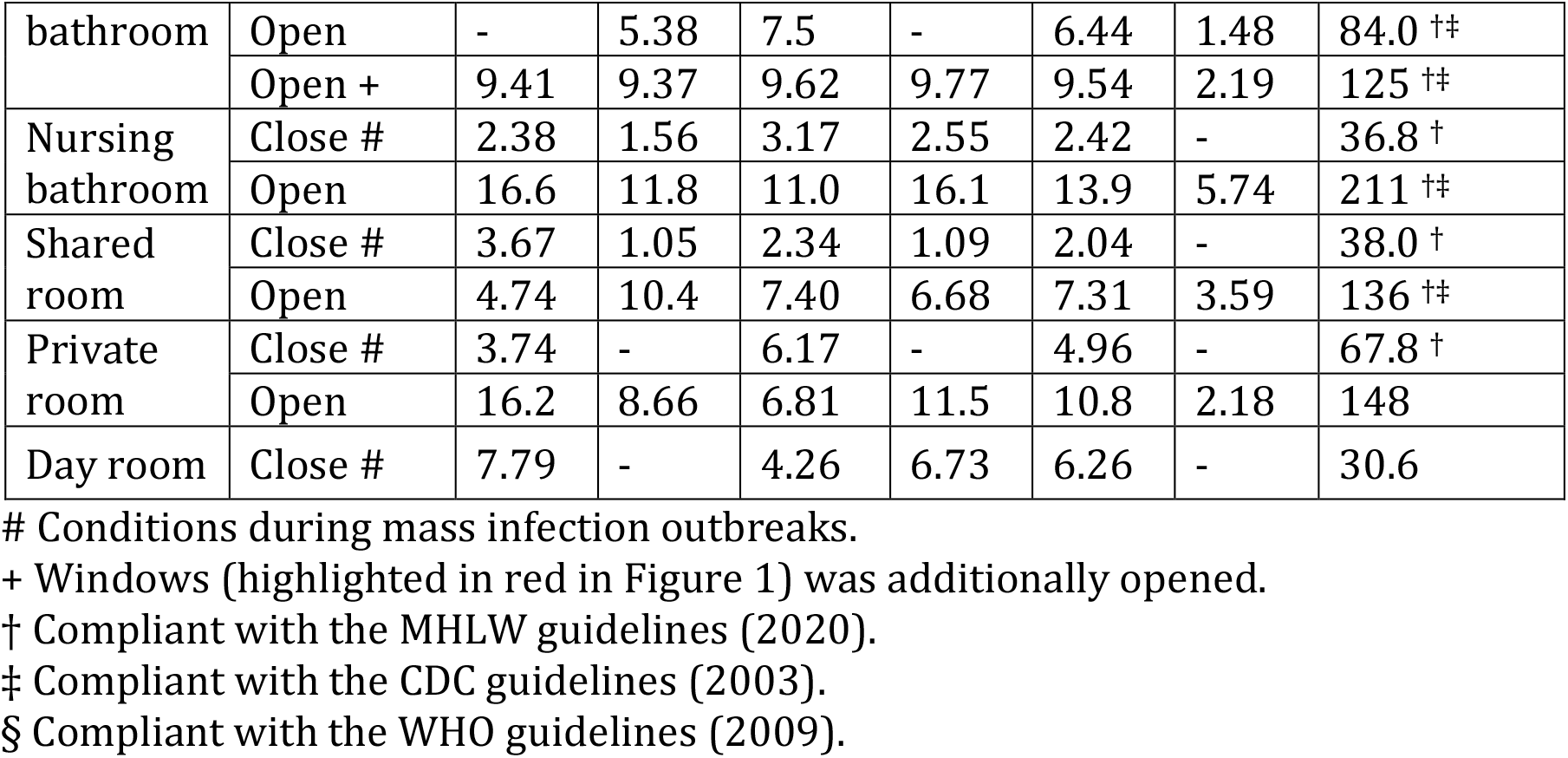
ACR values and per capita ventilation volumes calculated based on the primary measurement results in each room shown in Figure 1.

We conducted a factorial effect analysis using a linear regression with the estimated ACR as the objective variable and the window opening conditions, sensor location, and sensor model (P or K) as explanatory variables (Figures 4a‒4c). The results showed that ventilation was significantly dependent on the window opening conditions in all rooms (p-values of 0.0269, 0.00127, and 0.02838 for the regular bathroom, nursing bathroom, and shared room, respectively). In contrast, there was no significant dependence on the location of the sensor in any of the areas. Therefore, it is suggested that room ventilation was uniformly improved by opening the window in the regular and nursing bathrooms and shared room. There was no significant dependence on the sensor model.

**Figure 4.**
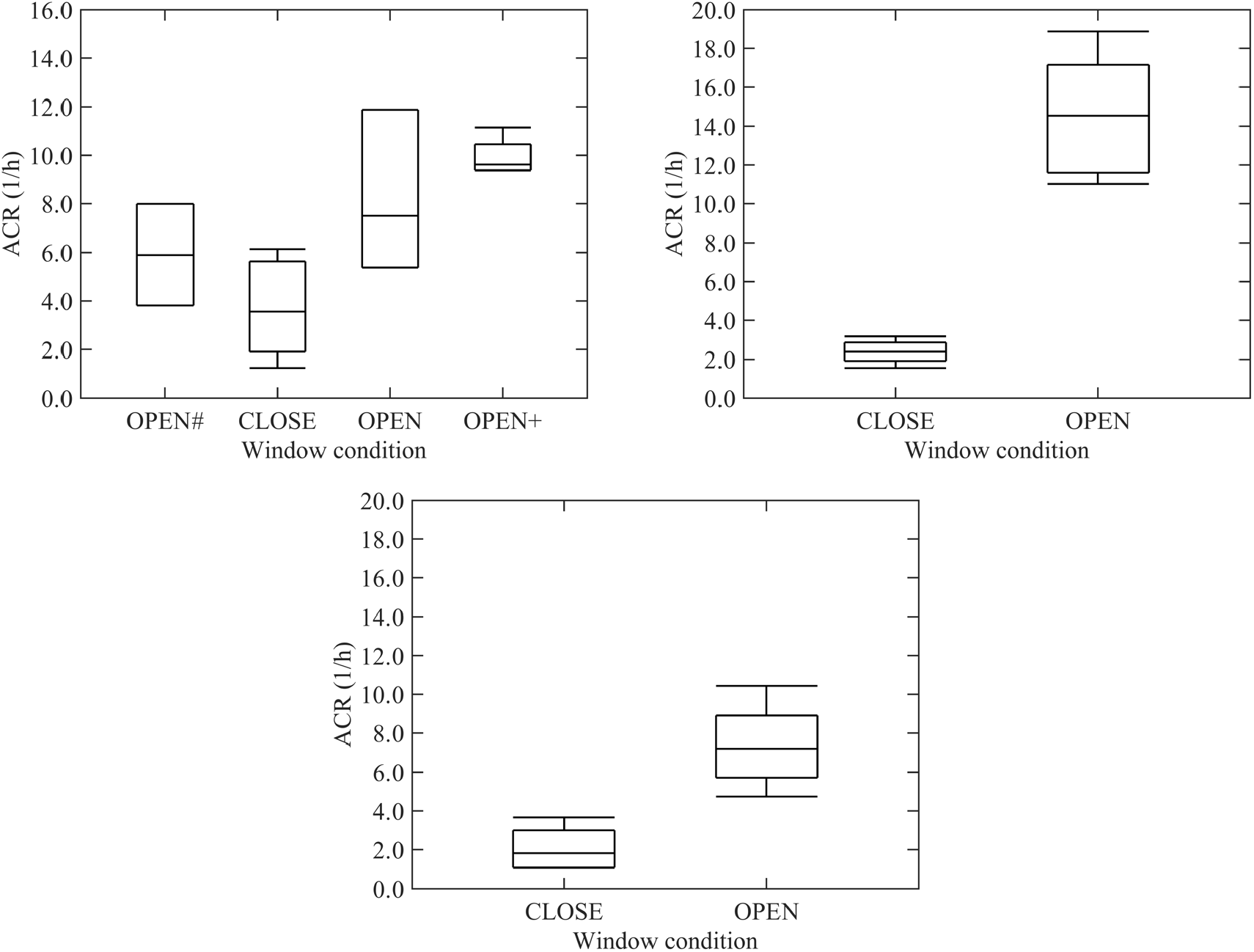
Factorial effect analysis using the linear regression model. The estimated ACR was used as the objective variable, and the presence/absence of window opening/closing, sensor location, and sensor model were used as explanatory variables. (a) Regular bathroom, (b) nursing bathroom, and (c) shared room. See Table 1 for the symbols # and +.

In the private room, the P and K2 sensors were installed close together because of the limited size of the room. Thus, we used a generalized linear mixed model to estimate and test the effects of ACR and window-opening conditions (with ventilation time and window opening set as fixed effects) and the difference between sensor models (with the sensor model as a variable effect). During the covariance parameter estimate of the variable effect, the Wald p-value of the sensor’s measurement was not significant (p-value of 0.9631). Therefore, no difference in the sensor’s measurement was confirmed. However, for the parameter estimates of fixed effects, the effects of ACR and window opening are both significant (p-value <0.0001). Therefore, for the private room, window opening uniformly improved the room ventilation.

The window opening conditions considered in the present experiments are also summarized in Table 1. During the mass infection outbreak in the facility, windows were closed, as marked with “#” in Table 1. Even under such window-closed conditions, all rooms met the MHLW’s standard for ventilation (>30 m^3^/h per person) [6], suggesting additional measures for ventilation were not necessary. Furthermore, the *r*_*ACR*_ values in Table 1 show that in all rooms, except the day room where the window-open experiment was not conducted, the ACR substantially improved by 1.48–5.74 times by opening the window to meet the CDC standard [4]. There was no significant dependence on the location or model of the sensor in all rooms where the window-open experiments were conducted, which suggests that the proposed method of estimating the ACR works robustly. Notably, even under the window-open conditions, no rooms met the WHO criteria [5]. However, as this facility is not classified as an infectious disease ward, this level of ventilation capacity is not strictly required.

Temporal variation of the measured CO_2_ concentrations in the wider area, including the private room and the day room (Figure 2), are shown in Figure 5. After the generation of CO_2_ gas was stopped at the step (4) discussed in the primary measurement section at *t* = *t*_0_, the concentration at sensor P steadily decreased with time. The CO_2_ concentration measured at K1 to K3 somewhat fluctuate; however, they generally show a gradual increase towards *t* − *t*_0_ = 9 (mins). The concentration at K3 is substantially higher than the other two K sensors from the early stage of the measurement (*t* − *t*_0_ = 2 mins), suggesting that the generated CO_2_ was predominantly advected from the private room by a large flow pattern, rather than simple diffusion process. This is also supported by the result discussed in Figure 4, which suggests the ventilation performance solely depended on the window opening condition, which dictates the air flow. Later in the measurement (*t* − *t*_0_ > 9 mins), the CO_2_ concentrations at K1 and K4 surpassed that of K3, with a relatively steep slope in the variation. Such rapid change in the rate of concentration increase was clearly due to advection rather than diffusion.

**Figure 5.**
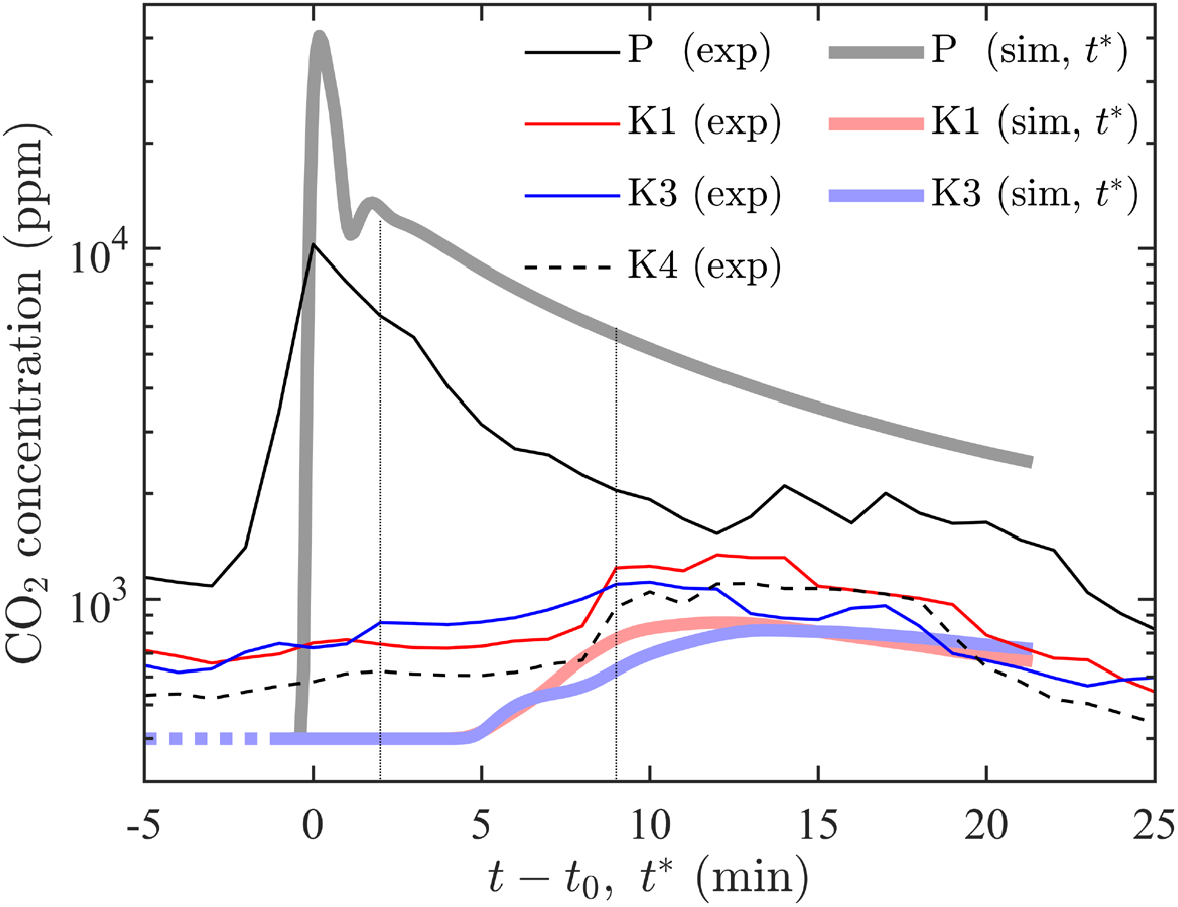
Temporal variation of CO2 concentration measured during the secondary measurement (thin line), and the numerical simulation (thick line) at the sensor positions shown in Figure 2. Note that the modified time axis *t*^*^ is used for the simulation result for comparison. The vertical lines indicate *t* − *t*_0_(= *t*^*^) = 2 and *t* − *t*_0_(= *t*^*^) = 9.

The temporal CO_2_ concentration evolutions are also shown in Figure 5 for the numerical simulation at the P, K1, and K3 probe positions. Despite attempts to mimic the environment in the nursing home facility during the experiment, some uncertainties in experimental setup, such as the initial field and exact boundary conditions for ventilation/air conditioning devices, are unavoidable. Although this causes discrepancy between the simulation results and measured data, the CO_2_ concentration obtained from the numerical simulation shows quantitatively similar levels to the measured values. Furthermore, the CO_2_ concentration evolution shows similar trends after the multiplication of the time axis of simulation results by a posteriori factor of 2.5, i.e., *t*^*^ = 2.5*t*_*sim*_ − *t*_*sim*,0_, where the subscript “sim” denotes the physical time in the simulation.

As for the evolution of the CO_2_ concentration at P simulated computationally, a peak is observed at *t*^*^ = 0, and its value is approximately 10^4^ ppm. Then, a monotonous decrease mode occurs until the end of the simulation. Such a mode transition is similar to the measurement result at the P sensor although the peak is four times larger than the measurement value, and its decrease is locally (*t*^*^ ≈ 1) nonmonotonic. The simulation and measurement results differ because of the uncertainties of the initial and boundary conditions in the simulation, mimicking the experimental setup. For example, the small gaps between the wall, door, and curtains, airflows through them, flow direction from air conditioning devices, and working staff in the test room cannot be accurately considered in the 3D model. However, the overall trend observed in the measurement is well reproduced in the numerical simulation.

Figure 6 shows instantaneous snapshots of the CO_2_ isosurface at 1000 ppm at *t*^*^ = 0.0, 0.9, 2.8, and 8.4 mins. A high CO_2_ concentration was observed in the entire private room when the CO_2_ generation was cut off at *t*^*^ = 0. Here, the maximum CO_2_ concentration was 8.8 × 10^5^ ppm. Owing to turbulent and molecular diffusion effects, the maximum CO_2_ concentration monotonically reduced to 2.9 × 10^5^ ppm (*t*^*^ = 0.9 mins, Figure 6b), 5.1 × 10^4^ ppm (*t*^*^ = 2.8 mins, Figure 6c), and 1.1 × 10^4^ ppm (*t*^*^ = 8.4 mins, Figure 6d). Furthermore, the region with a CO_2_ concentration >1000 ppm (high-concentration zone), which is substantially greater than the background value, *C*_0_, spread across the day room and corridor. In particular, the high concentration zone occupied the corridor in front of the private room at *t*^*^ = 2.8 mins. The local fluid velocity, overlaid on the CO_2_ isosurface, shows that the leading edge of the CO_2_ isosurface moving towards the day room (see red arrows in Figures 6b and 6c) yields a relatively large velocity. Therefore, the large-scale flow dictated the spread of the gas mixture containing infectious aerosols rather than the molecular diffusion process.

**Figure 6.**
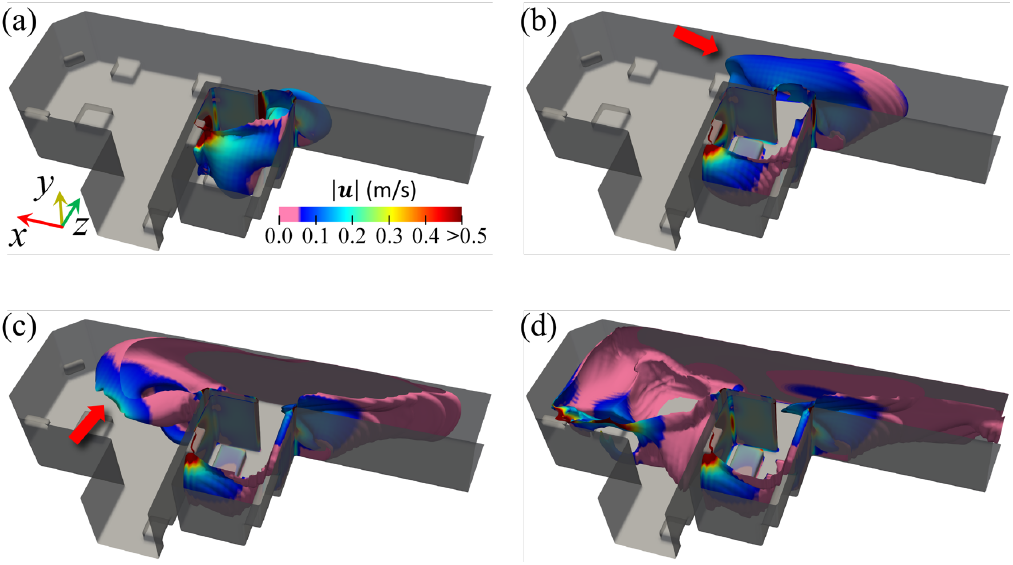
Temporal evolution of isosurface of CO_2_ concentration at 1000 ppm at *t*^*^ = 0.0, 0.9, 2.8, and 8.4 mins. The local color of the isosurface shows local velocity magnitude in m/s, (rainbow: high-velocity region, pink: low-velocity region).

## Discussion

Based on the present analyses, by opening windows, which is a low-cost method, to significantly improve ventilation can better suppress aerosol infection in elderly care facilities. Furthermore, in this facility, both regular and nursing bathrooms were equipped with jalousie windows to ensure the privacy of the residents. Thus, opening windows for additional ventilation had no negative impacts on their privacy.

However, in many elderly care facilities, opening windows is avoided. This is because of concerns, amongst facility staff and residents’ family members, regarding dangerous behavior, such as escaping or jumping out of the opened windows due to the behavioral and psychological symptoms of dementia (BPSD), which some care home residents may have. Therefore, opening windows as a measure to enhance room air ventilation should be carried out while considering these safety aspects. For example, a simple locking mechanism can be installed to prevent windows from opening only at a certain width, thus effectively ensuring safety and ventilation. Additionally, future studies should clarify the effect of opening windows on heating and cooling.

Moreover, results of the secondary measurement discussed in Figures 5 and 6 suggest that the aerosols can be advected between different rooms by the relatively strong fluid flows created by the ventilation fans and air conditioning devices. Generally, the required ventilation volume (per hour) is determined by the capacity of the room. For smaller rooms, such as the private or shared room in the present study, the required ventilation volume tends to be smaller, and vice versa for larger rooms, such as the day room. Therefore, in open or semi-open buildings where multiple rooms are spatially connected, such as the present facility, advective flows from private to common areas may exist, and adhering to the ventilation volume standard for each room individually may not be sufficient to prevent mass infection outbreak such as in the present case. This is a challenge posed for most nursing homes, where private and common spaces are directly or indirectly connected.

Hunziker defines a specific form of propagation called “jet lidar” as a mode of transport of infectious aerosols that causes secondary infections of COVID-19 in hospitals: Jet lidar initially travels as an air jet with turbulence, and then falls away at a distance [8]. In future studies, we can improve the reproducibility of the infection situation in computer simulations by considering the jet rider produced by the infected person coughing.

Anderson et al. determined that typical elderly care facilities may be vulnerable to COVID-19 infections since they are designed to promote social interaction and collaboration among the residents [9] via common spaces (e.g., day rooms and areas for social activities) and hallways without partitions [10, 11]. This aspect is important for the residents’ social interactions and for daily monitoring by staff. In addition, in Japan, the deregulation of the Law for Partial Revision of the Building Standards Law (enacted on September 25, 2018), which exempted the floor area of common corridors from the calculation of the floor area ratio for nursing homes, may have provided an impetus for the active use of corridors as common relaxation areas. However, from an infection control perspective, there is room for improvement in these open-plan architectural guidelines. With these precedents, a practical guideline should be formulated specifically to address the operational patterns of elderly care facilities.

As an example of a temporary guideline during a pandemic, downwind transmission can be easily prevented by discontinuing the use of private rooms close to common rooms in elderly care facilities. The excessive installation of vinyl partitions may also contribute to mass infection [2] because of the stagnation of fluid flow essential for active ventilation. Therefore, care should be taken when designing partitions so that they do not interfere with ventilation. A more quantitative measure would be recommended to check the pressure difference between the room and hallway [4, 12], as recommended in healthcare settings. If the possibility of downwind transmission created by the pressure difference becomes apparent, some measures (e.g., transparent partitions or air curtains) can be taken to prevent the large-scale air flow, such as the one observed in the present physical or fluid-dynamic numerical simulation. These measures must consider the accessibility and visibility to ensure the quality of life (QoL) of residents. Another effective and less intrusive measure could be the use of High Efficiency Particulate Air Filter (HEPA) filters for purification of the airflow passing through.

## Conclusions

In this study, a real-world mass infection outbreak which occurred in an elderly care facility was simulated experimentally and numerically to investigate the controlling factors and quantify the effectiveness of various natural ventilation settings using the ACR. Using the CO_2_ tracer gas method, we determined that the low-cost intervention of opening windows can improve the ventilation frequency by a factor of 1.48–5.74. This implies that advective fluid flows are key to controlling the spread of high CO_2_ concentration zones.

A numerical simulation was performed to obtain spatio-temporal evolution on such high CO_2_ concentration zones under similar conditions to the present experiment. Development of high CO_2_ concentration zones occurs in the first few minutes. Furthermore, the leading edge of such zones towards the day room, where multiple residents gather for activities, yields relatively high fluid velocity, suggesting that the large-scale advective flow dictates the spread of such high CO_2_ concentration zones. The present results suggest that secondary infections could occur because of the topology of the aerosol advection driven large-scale flows, even if the ventilation is sufficient. Furthermore, this phenomenon may be influenced by the architectural design specific to typical elderly care facilities.

To prevent or deter the mass infection outbreaks in elderly care facilities, policies for guidelines on the architectural design and reviews of related laws will be necessary considering both the QoL of the residents and suppression of large-scale flow towards communal areas. In addition, quantitative studies and interventions are required to avoid downwind contamination in existing buildings.

## Data Availability

All data produced in the present study are available upon reasonable request to the authors

## Acknowledgements

This work was supported by JSPS KAKENHI Grant No. 21K19820 and KDDI foundation.

## Authors’ Contributions

YI conceptualized and designed the study and contributed manuscript writing, and review and approval of the paper. SY performed formal analysis, manuscript writing and review and approval of the paper. YM performed data visualization and manuscript writing, and review and approval of the paper. AS, HK and YK contributed data extraction and investigation. All authors approved the final manuscript as submitted.

## Conflicts of Interest

The authors have no potential conflicts of interest to declare.

### Abbreviations

ACR: air change rate per hour
CDC: Centers for Disease Control and Prevention
MHLW: Ministry of Health, Labor and Welfare
WHO: World Health Organization

## Notes

**Data availability statement:** The data that support the findings of this study are available from the corresponding author, Y.I., upon reasonable request.

### Competing Interest Statement

The authors have declared no competing interest.

### Author Declarations

This study was approved by the Ethics Committee on Experiments on Human Subjects in the Corresponding author's institution (the University of Electro-Communications, Tokyo, Japan). The approval number is 21005.

